# Non-Nutritive Suckling System for Real-Time Characterization of Intraoral Vacuum Profile in Full Term Neonates

**DOI:** 10.1101/2022.12.14.22283473

**Authors:** Phuong Truong, Erin Walsh, Vanessa P. Scott, Todd Coleman, Gopesh Tilvawala, James Friend

## Abstract

Infant breastfeeding diagnostics remain subjective due to the absence of instrumentation to objectively measure and understand infant oral motor skills and suckling characteristics. Qualitative diagnostic exams, such as the digital suck assessment which relies upon a clinician’s gloved finger inserted into the infant’s mouth, produce a diversity of diagnoses and intervention pathways due to their subjective nature. In this paper, we report on the design of a non-nutritive suckling (NNS) system which quantifies and analyzes quantitative intraoral vacuum and sucking patterns of full-term neonates in real time. In our study, we evaluate thirty neonate suckling profiles to demonstrate the technical and clinical feasibility of the system. We successfully extract the mean suck vacuum, maximum suck vacuum, frequency, burst duration, number of sucks per burst, number of sucks per minute, and number of bursts per minute. In addition, we highlight the discovery of three intraoral vacuum profile shapes that are found to be correlated to different levels of suckling characteristics. These results establish a framework for future studies to evaluate oromotor dysfunction that affect the appearance of these signals based on established normal profiles. Ultimately, with the ability to easily and quickly capture intraoral vacuum data, clinicians can more accurately perform suckling assessments to provide timely intervention and assist mothers and infants towards successful breastfeeding outcomes.

## I. INTRODUCTION

Breastfeeding is a natural biologic function that fosters attachment and safeguards the health of mothers and babies. By breastfeeding, mothers experience lower risks of reproductive organ cancers, type II diabetes, cardiovascular disease and mental health disorders, while infants experience lower risks of infectious diseases, gastrointestinal and respiratory health issues, allergies, type II diabetes, hypertension, and obesity [1–5]. While the advantages of breast milk far outweigh formula, the rate of exclusive breastfeeding at six months post-birth plummets to only 25%, according to the CDC’s 2018-2019 National Immunization Survey [6].

Early breastfeeding diagnostics to identify poor latch and suck are essential for timely interventions and support for the mother and infant to help reduce breastfeeding cessation. Presently, feeding clinicians and pediatricians assist mothers and infants with breastfeeding challenges, yet are constrained by the absence of instrumentation to objectively quantify suck vacuum, a key aspect of successful breastfeeding [7–9]. Existing assessment methods are essentially qualitative measurements, such as digital suck assessment using a gloved finger to determine infant suckling vacuum [10]. While more elaborate assessment scales do exist, few clinicians are trained to administer and interpret them. Due to this, both objectivity and consensus among the clinical community are lacking [11], leaving the diagnosis of breastfeeding difficulties in an ambiguous limbo and resulting in a variety of interventions that may be unwarranted (e.g. frenotomy). These difficult circumstances ultimately causes infants to undergo unnecessary surgery, putting them at risk of bleeding, pain, infection, ulceration, and other complications [12, 13].

In recent years, several devices and systems have been developed to quantify the suckling profile and oral-motor coordination of premature infants [14–30]. These systems principally address the challenge of oral feeding readiness in premature infants by measuring their intraoral suction (vacuum) and expression (contact) pressure. While not posed to diagnose breastfeeding problems in full-term infants, some of these systems show promise in doing so. Grassi, et al., for example, developed a sensorized pacifier that measures suction and expression pressures using two integrated pressure transducers, displaying measurement results via a simple graphical user interface (GUI) [14]. Lau, et al., studied pressure measurements from two sensorized catheters attached to a gloved index finger [22, 25]. Ebrahimi, et al., devised a portable compact intraoral pressure measurement system that includes features such as a custom printed circuit board, wireless communication, and a rechargeable battery [18]. The FDA-approved NTrainer system measures the displacement of the tongue (expression pressure) and incorporates pneumatic actuation to help facilitate infant oromotor skills [31–33]. Geddes et al., utilizes ultrasound along with pressure transducers to correlate vacuum characteristics to milk intake during nutritive sucking [7]. These devices, along with many others [14–25] proposed in the literature, all reflect an effort to provide objective quantification of infant intraoral vacuum.

Despite many studies addressing training and coordination of non-nutritive suck in premature infants, very few in the literature have emphasized the development of instrumentation aimed to assess healthy newborn infants experiencing breastfeeding difficulties. While technologies of similar function and purpose dating back over two decades do exist, they have not yet emerged to change medical practices due to their problems with clinical adoptability and measurement reproducibility [22, 34, 35]. As a result, subjective metrics to identify oral dysfunction, such as ankyloglossia, remain widespread and controversial in the clinical community while breastfeeding rates remain low [11]. As intraoral vacuum is well recognized to play a key part in infant suckling and milk removal, our aim is to address the need for screening instrumentation to assess infant non-nutritive suckling (NNS) vacuum) [36].

In this paper, we report on the design of a non-nutritive suckling (NNS) system to measure and analyze intraoral vacuum of full-term neonates in real-time. Our system considers factors important in translation to clinical use, including real-time analysis with immediate feedback to the clinician, ease of use, measurement accuracy and repeatability, and accounting for variability in infant suckling preferences. Our system design provides an objective alternative to the standard digital suck assessment. Specifically, we measure in full term infants the suckling vacuum to extract the following objective micro-structure parameters: the mean and maximum vacuum amplitude, suckling frequency, number of suckling events per burst, burst duration, and number of bursts per minute. Our findings show that the infants’ intraoral profile produce distinctive vacuum responses that can in turn be used to identify orofacial issues. We categorize these signals and provide a framework for studying oromotor dysfunctions in future studies.

## II. MATERIALS AND METHODS

### A. Clinical and Technical Requirements

To develop a robust system that is feasible for clinical use, our design approach for the NNS system considers its utilization and interaction with both clinicians and infants. Key parameters of the sensing system, described in Table I, and the configuration of the components were considered as a part of the design of the NNS system to ensure clinical feasibility. Table I also summarizes the design requirements for our proposed system based on the advantages and drawbacks of existing systems reported in the literature.

**TABLE I.** Design parameters considered in the NNS system to meet clinical and engineering requirements needed for clinical feasibility.

| Design Requirements |  |
| --- | --- |
| Ease of use | Hardware and software must be intuitive for clinicians to use and manage with minimal training |
| Biosafety | Components in direct contact with saliva, bodily fluids and oral cavity must be sterilized before use and must be sterilized or disposed after each use |
| Adaptability | Infant pacifier preferences may vary; suckling unit must be versatile in adapting to various pacifier types |
| Biocompatibility | Components interfacing with infant must meet biocompatibility safety requirements |
| Electrical Safety | Electrical components must operate within International Electrotechnical Commission (IEC) safety limits |
| Accuracy | Pressure sensing unit dynamic range must be able to measure physiological range of intraoral vacuum of infants (0 mmHg to $-400$ mmHg) |
| Repeatability | System measurements must be repeatable as needed to track infant vacuum over time |

### B. NNS System Hardware

To achieve these design requirements, we developed an NNS system that is comprised of four main components: a single-use modified pacifier, a pressure sensor, a data acquisition unit, and a custom-made software interface (Fig. 1).

**FIG. 1.**
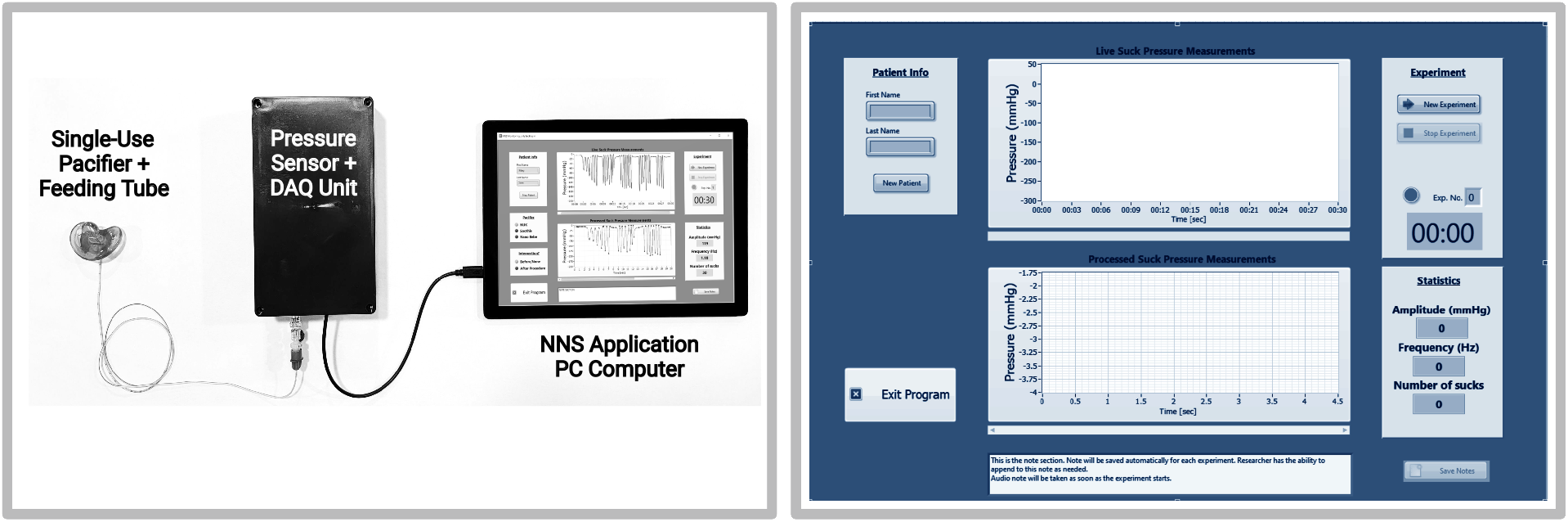
Image of the NNS system design with four major components: a modified pacifier, pressure sensor, data acquisition board, and a custom software interface. The design considers the intended clinical use and ease of adopting the system. The system can be used with minimal training and seamlessly integrated into the clinical workflow.

The pacifier component was fabricated using a commercial teat (Orthodontic Pacifier, NUK) integrated with a 36-inch 5 fr non-collapsible feeding tube (Kangaroo Neonatal & Pediatric Feeding Tube, Covidien). The air in the tubing has significantly affected sensitivity of other devices in the literature that use large volume tubing. We utilize a very narrow tubing (5 fr outer diameter, 1 mm inner diameter) to reduce the total volume of air that can be compressed in the system. This helps us avoid the adverse vacuum measurements seen in other devices: it minimizes the air volume but is large enough to avoid boundary layer losses and drag. Furthermore, 36-inch tubing was the desired length to provide sufficient slack length for clinician and infant during measurements. While this teat was selected for its shape and fit with the infants’ oral anatomy [37], the modularity of the system permits quick substitution with any pacifier shape and type preferred by the infant. To integrate the feeding tube with pacifier, a 1-mm diameter biopsy punch was used to create an opening at the tip of the pacifier and the feeding tube was passed through the opening. Next, 0.1 mL of polydimethylsiloxane (Sylgard 184, Dow Corning), a biocompatible and inert non-toxic silicone was used to hold the feeding tube in position at the pacifier’s tip. The silicone was mixed at a 20:1 ratio to produce a nearly gel-like elastic material to mimic the pacifier material, and the volume used was the minimum amount required to hold the feeding tube in place at the tip, leaving the majority of the pacifier and its tip empty. This helped us avoid altering the original stiffness characteristics of the pacifier. The silicone was cured in a 50°C oven for 8 hours. Once integrated, the modified pacifier was cleaned with water and mild soap and dried. The unit was bagged and sterilized under 275 nm ultraviolet light (Sterilizer and Dryer, VANELC) for 35 minutes. The bio-compatibility and safety of the modified pacifier was considered in the design. We limit infant exposure to any unknown materials and only consider those that are accepted or widely used. A silicone pacifier (commercially available) integrated with a medical-grade PVC feeding tube are the only materials in contact with the infant, ensuring biocompatibility and safety. In circumstances where the infant rejects the pacifier or has a known allergic reaction to the pacifier material, the pacifier can be substituted for any preferred pacifier such as the Soothie (Philips AVENT, Tucson, AZ), a standard pacifier used in hospitals.

A piezoresistive pressure sensor (MPX5100AP, NXP Semiconductors, Eindhoven, Netherlands) was selected with an operating range of 110–860 mmHg (absolute) to fit the application and system design requirements of neonatal suckling dynamic range. The intraoral vacuum of typical neonates during suckling has been reported to be 375–825 mmHg [9, 14]. The sensor was calibrated against a pressure gauge at various vacuum conditions to verify the manufacturer’s reported specifications [38]. Once we verified its accuracy and repeatability, the sensor was electronically configured to begin measurements. To acquire intraoral vacuum measurements, the pressure sensor was directly connected to the modified pacifier and feeding tube. A data acquisition board (myDAQ, National Instruments, Austin, TX) collected the pressure measurements and was sent to a computer with a graphical software interface (LabVIEW, National Instruments) for simple analysis and data visualization by the clinician. The sampling frequency was set to 1000 Hz to sample at a greater rate than the suckling frequency, which is reported in the literature to be within the 1.5 Hz to 2.5 Hz range [14, 39]. The maximum output voltage in the device is 5 VDC, well below the standard limit for contact with a human (∼ 30 VDC). Moreover, the maximum current available in the device is about 2 mA. These aspects make the device intrinsically safe according to IEC standards 61140, 60364, 61010-1, and 60479. The data acquisition board and sensor were entirely contained in an insulated box without possibility of making contact with the infant.

The pacifier and feeding tube unit connected to the pressure sensor through a quick connect luer lock allowing for ease of use. The design considers the clinical workflow as follows. To use the unit, a clinician would (1) connect the hardware to a computer via USB, (2) open the NNS software, (3) open a new pacifier unit, (4) connect the pacifier tubing to the hardware, and (5) press `start experiment` to collect data. All of this can be done in less than a minute with minimal training. Finally, since the components are relatively low cost, we designed the system such that the pacifier-feeding tube unit is single-use (disposable) to minimize both cross-contamination of fluids such as saliva between patients and the need to clean or sterilize the device after each measurement. The disposability and quick connect/disconnect design features helps facilitate the integration of the device into the fast-paced clinical workflow and allows the clinician to quickly test patients as a part of their routine examination schedules.

### C. System Software Design and Signal Processing

The NNS system software is designed to record, process, and display intraoral vacuum measurements for the clinician to see while the data collection is underway. This allows clinicians to utilize information for rapid diagnosis and dynamically adjust to retake measurements as needed. Table II summarizes the key software features that enable rapid diagnosis in a clinical setting.

**TABLE II.**
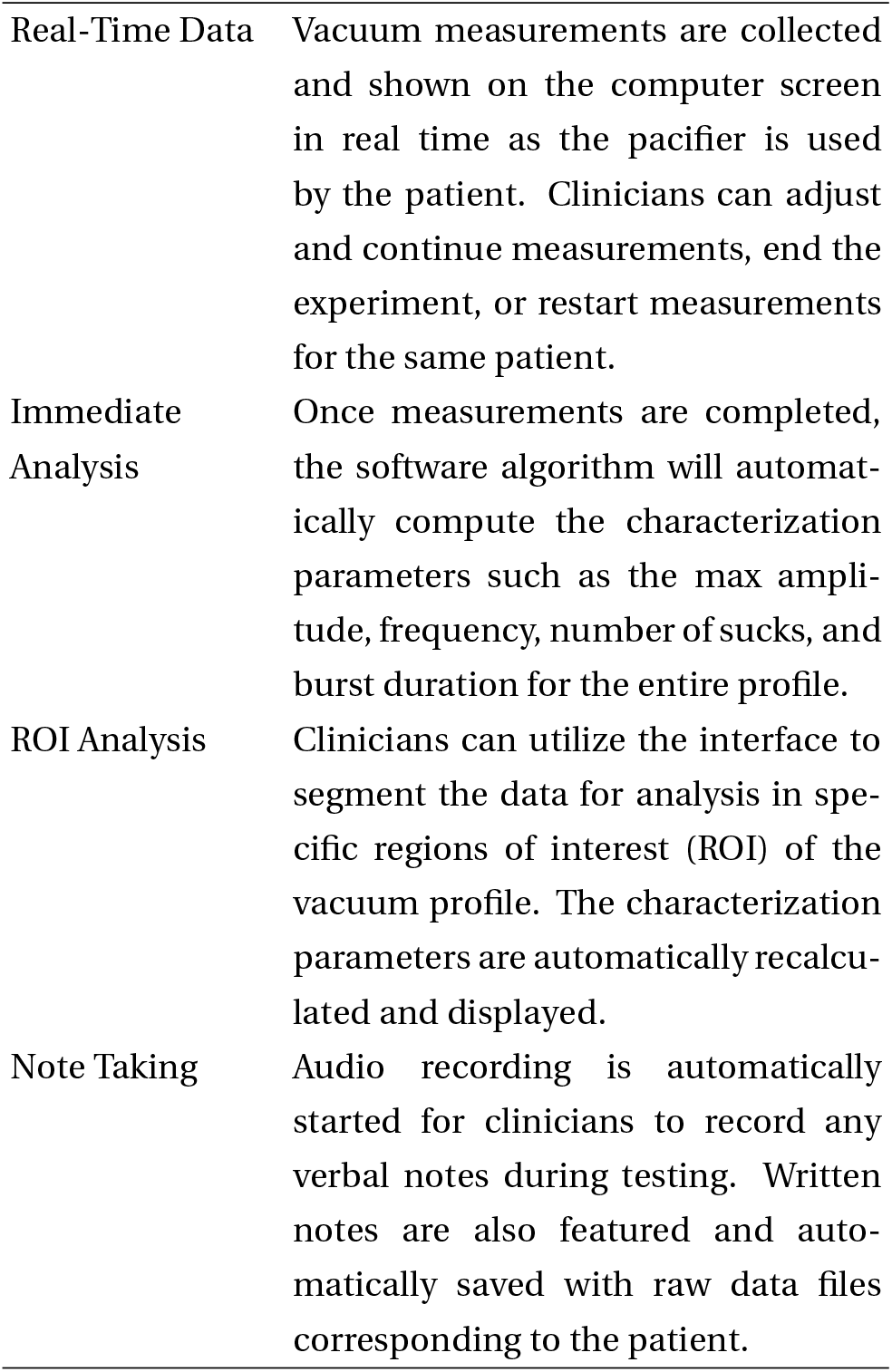
Software features and capabilities of the NNS application. Its design focuses on the clinical needs of the medical professional in a clinical breastfeeding assistance setting.

The NNS application was designed and built using LabVIEW, a graphical programming environment. The custom program was packaged into an executable application that can be deployed on any PC that is readily available in the clinic without the need of the native LabVIEW software. This allows for ease of adoption and reduces barriers to entry. The NNS app was designed with an intuitive user interface where the clinicians can enter patient information, start (or stop) experiments, and view the pressure profile and key metrics in real time. Clinicians may also magnify regions of interest for closer inspection and analysis of the shape of the suckling signal. Region of interest analysis is an important feature due to the unpredictability of infant behavior that may be disruptive during vacuum measurements. To isolate abnormalities caused by disruptions, clinicians can perform analysis on specified regions immediately after a test.

The software begins by calibrating the sensor to remove any baseline drift caused by the sensor. The clinician proceeds to insert the pacifier in the infants mouth to collect NNS data. Should there be difficulty, the clinician can restart the measurement as desired. Once the signal is acquired, characterization is performed automatically by the software. Table III describes the parameters extracted from the NNS signal. The analysis sequence of the app begins with the detection of peaks and valleys for the full suckling profile. Referring to Fig. 2, one suck cycle is defined as having a minimum suck amplitude of 10 mmHg based on reported definitions in the literature [40]. These values are used to establish the threshold of the peak-valley detection algorithm. A burst is defined as two or more consecutive suck cycles with a minimum rest period of one second between bursts [41]. From the NNS profile, other characteristics can be extracted. The suck amplitude is defined by the average measured amplitude of the infant’s vacuum placed upon the pacifier over the trial. The time period between two successive valleys of locally maximum suck vacuum are collected over all the suckling events and used to calculate the average suck frequency, both for each burst and for the entire trial.

**TABLE III.**
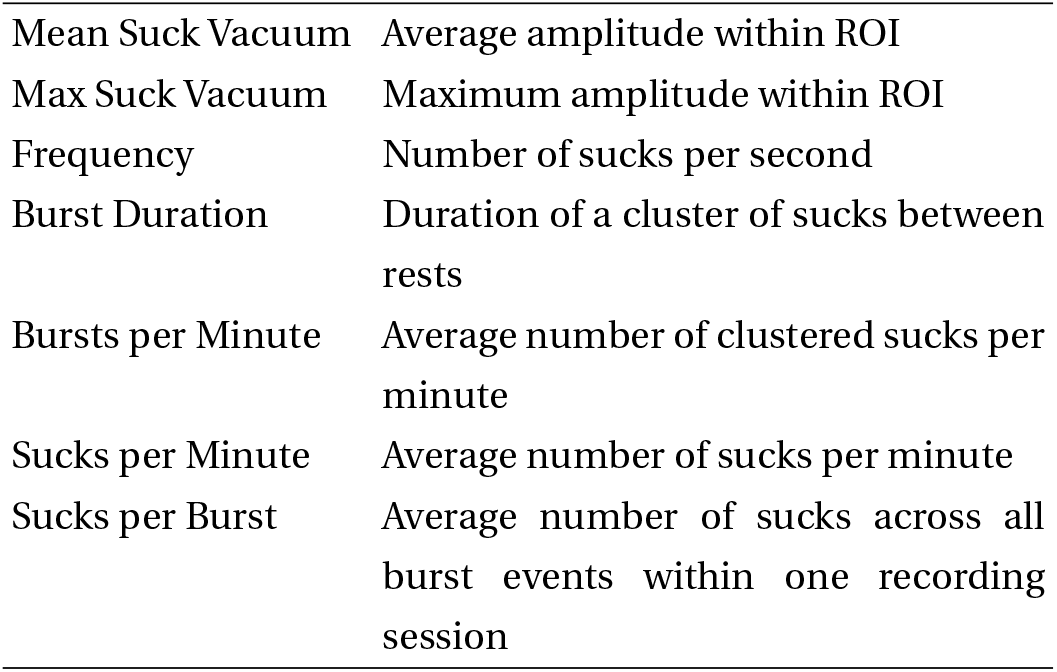
The features extracted from the suckling signal by the NNS software and which help to characterize the infant’s suckling.

**FIG. 2.**
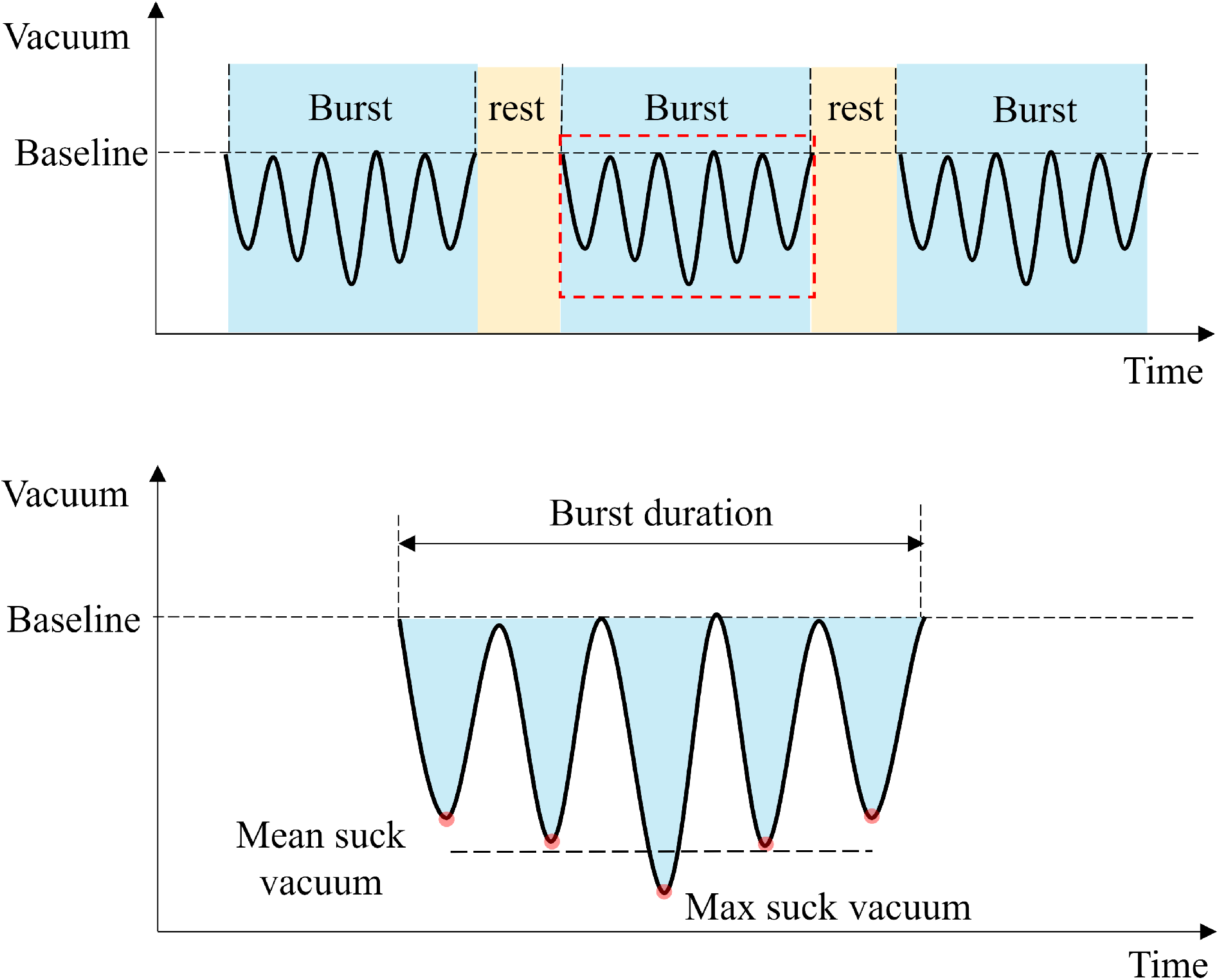
Illustration of a typical intraoral vacuum waveform labeled with characterization parameters.

An interactive cursor allows the clinician to extract these features for a specific region of interest (ROI). The application automatically updates values as the clinician selects different ranges of the measured data via the GUI. This enables the clinician to focus on specific time points or ROIs in the suck profile for a closer analysis.

### D. Clinical Testing and Protocol

Thirty healthy term newborns (gestational age: 37–42 weeks) under 30 days of age were recruited from both the UC San Diego Health Department of Otolarngology’s Center for Voice and Swallowing and the Pediatrics Department. The infant inclusion criteria for the study were: (1) infants 4–30 days old (critical period to establish breastfeeding); (2) healthy; no significant birth or post-partum complications; and (3) no known allergy to silicone or elastomers typically used for pacifiers and bottle nipples. The study aimed to measure infant suckling vacuum using the NNS system to establish a norm for sucking signal characteristics. Testing occurred during a lactation consultation visit (Center for Voice and Swallowing) and during outpatient pediatric visits (UC San Diego General Pediatrics). Approval from the Institutional Review Board at UC San Diego (IRB 800070 approved 13 September 2021) was obtained before recruitment started. Parents were informed of the nature of the study and consented before the experiment began. Infants underwent a routine weight and physical exam. After an initial routine evaluation, infants were offered the NNS system pacifier. Before the start of measurements, the infant’s seal around the pacifier was verified to be secured and established. Inadequate seal can be observed through the infant’s contact with pacifier and an observable abrupt loss in vacuum. If the seal was determined to be inadequate, the measurements were repeated. The intraoral vacuum was recorded for a duration of 60 seconds.

## III. RESULTS

Figure 1 shows the final and complete NNS system. The results from the clinical study validate the ability of the NNS system to measure intraoral vacuum. Clinicians utilized the system with minimal training and were able to incorporate the system into their workflow. The characteristics described in Table III were collected over 60 seconds. Figure 3 is a representative snapshot of a typical infant intraoral vacuum profile, including the details of a particular burst event. Table IV summarizes the parameters extracted from the cohort of 30 infants’ suckling data. Values extracted are comparable to those previously reported in the literature, both during NNS [14, 35, 42] and breastfeeding [43] as shown in the table, demonstrating the systems’ ability to capture intraoral vacuum over time.

**FIG. 3.**
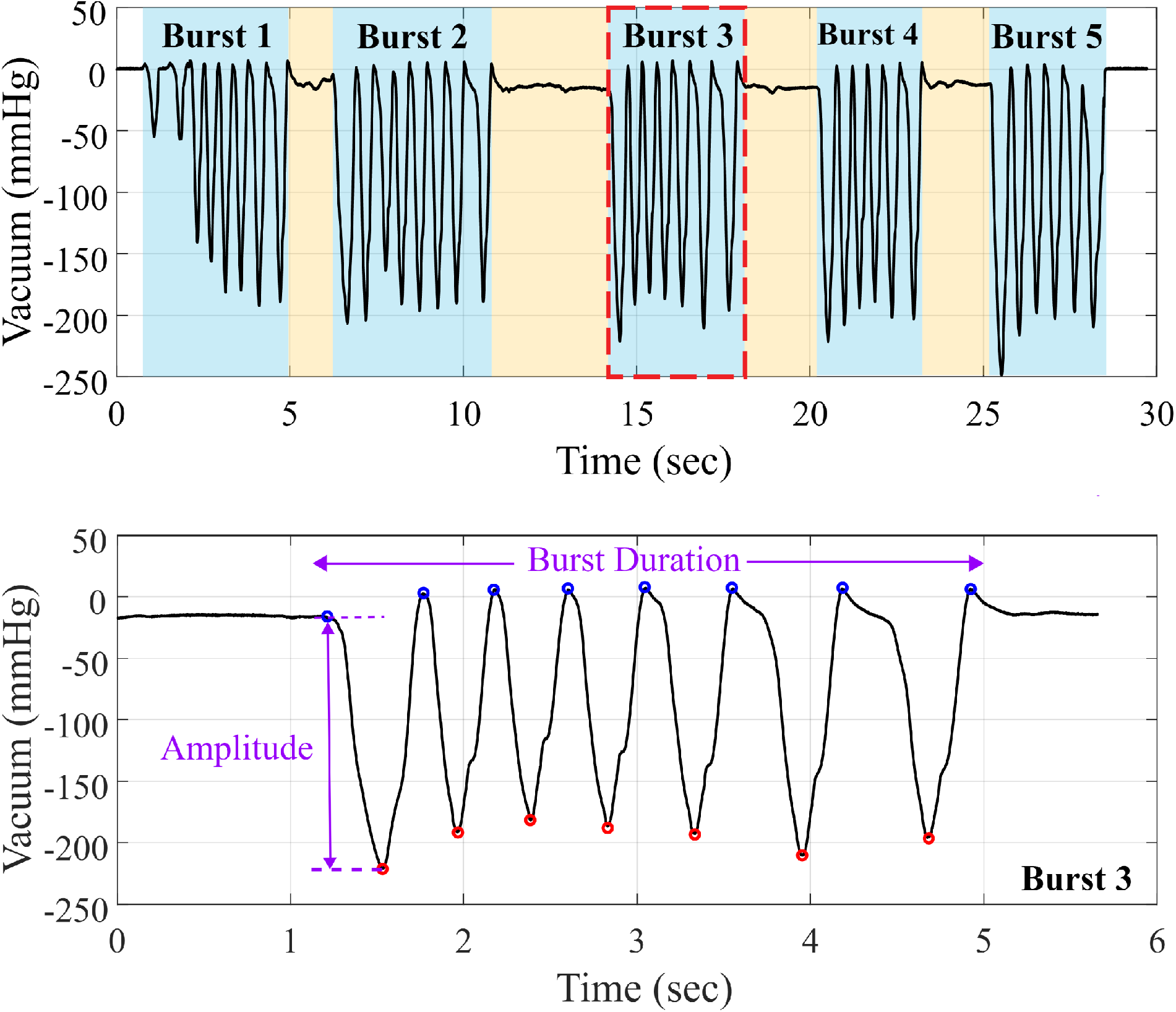
An example of a suckling signal generated by an infant utilizing the modified pacifier (left). The figure on the right shows an example of a region of interest (a zoomed in of Burst 3) generated from the NNS software, providing immediate analysis of the signal within the region of interest defined by the clinician. Analysis includes peak and valley detection to characterize the signal parameters such as burst duration, maximum suck vacuum, and mean suck vacuum.

**TABLE IV.** A summary of results comparing extracted parameter values collected in this study and those reported in the literature.

| Parameters | This Work | Grassi [14] | Zimmerman [42] | Hafstrom [35] | Prieto [43] |
| --- | --- | --- | --- | --- | --- |
| Number of subjects | 30 | 9 | 16 | 53 | 17 |
| Mean suck vacuum (mmHg) | 118.6 (30.8) | - | 64.6 (22.9) | - | 50 (5.7) |
| Max suck vacuum (mmHg) | 143.6 (32.2) | 164.1 (38.6) | - | - | 197 (10) |
| Frequency (Hz) | 2.01 (0.37) | - | 2.16 (0.35) | 1.91 (0.27) | - |
| Sucks per burst | 8.8 (5.5) | 6.9 (1.0) | 5.6 (3.1) | - | - |
| Burst duration (sec) | 4.4 (3.0) | 2.9 (0.6) | 2.5 (1.4) | 4.0 (1.9) | - |
| Sucks per minute | 70.7 (16.9) | - | 28.1 (25.6) | - | 52 (26) |
| Bursts per minute | 7.2 (3.1) | 9.3 (2.1) | 4.1 (2.7) | - | - |

Upon closer inspection of the suckling signals by magnifying a suckling burst, we observe subtle differences in the vacuum transducer’s signal. Figure 4 shows signals representative of the three distinguishable profiles found in the NNS signal of the thirty infants tested. We classify the three shapes as: smooth sinusoidal, sharp valley, and double valley. While the factors contributing to these varying shapes are not yet known, we group the cohort of infant profiles into three groups corresponding each of the three shapes. Figures 5 and 6 graphically illustrate the statistical differences between each group based on their characteristics.

**FIG. 4.**
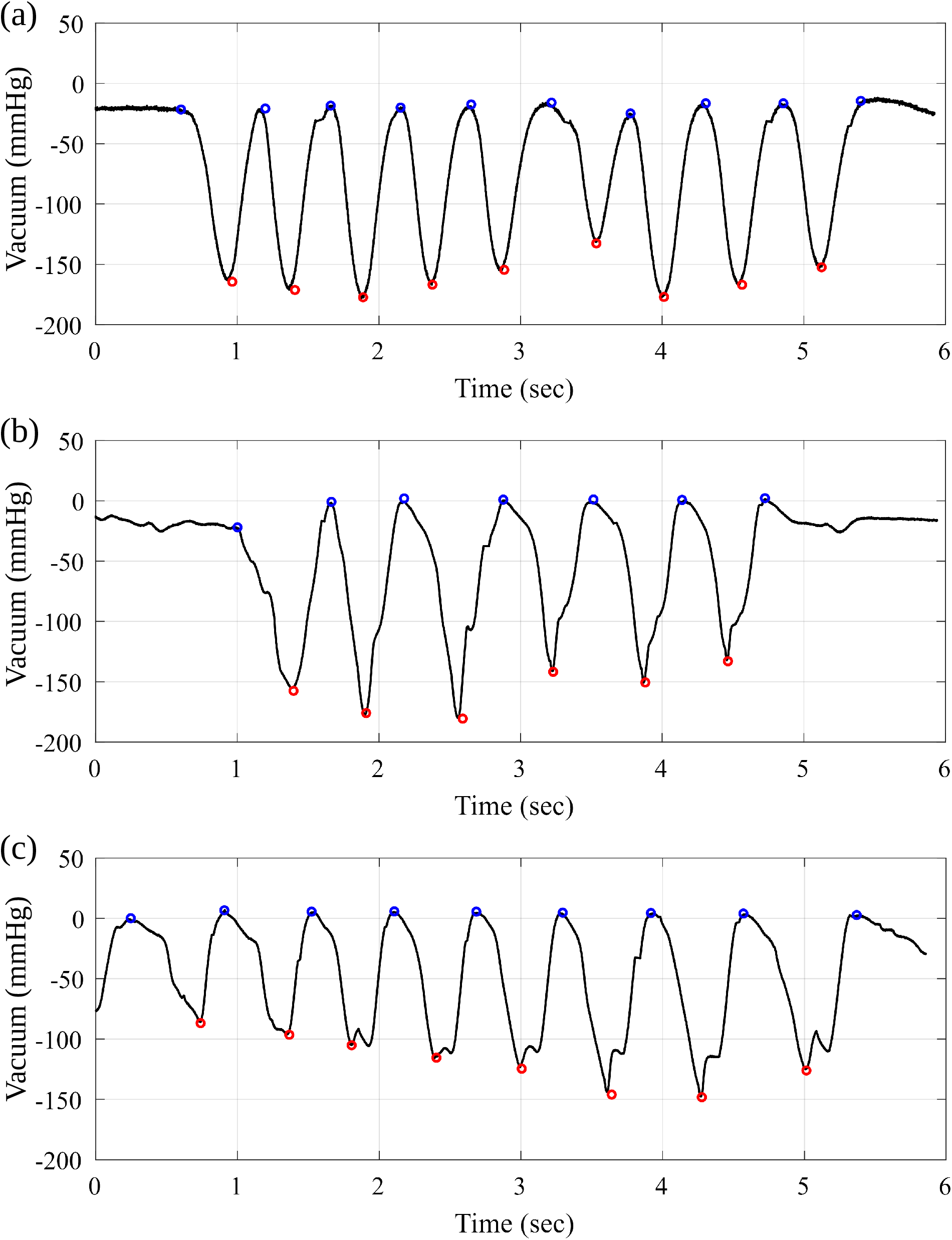
Three distinguishable profiles appear in the NNS suckling signal of 30 infants: (a) group 1: smooth sinusoidal, (b) group 2: sharp valley, and (c) group 3: double valley. This figure shows representative examples of NNS signal from each group.

**FIG. 5.**
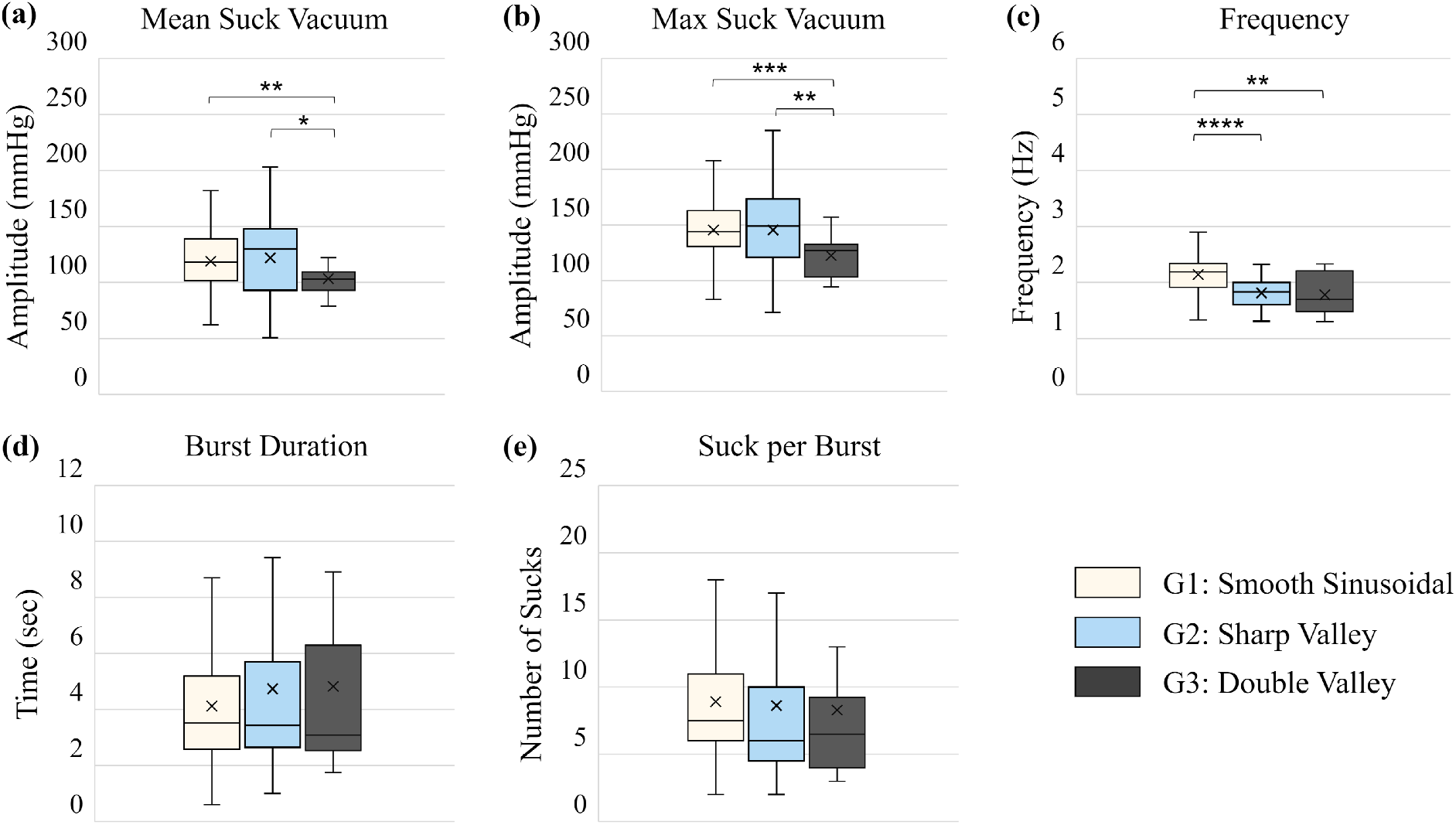
Box and whisker plots comparing the extracted parameters from three classified groups. We observe statistically significant differences between groups 1 and 3 across mean suck vacuum, maximum suck vacuum, and frequency.

**FIG. 6.**
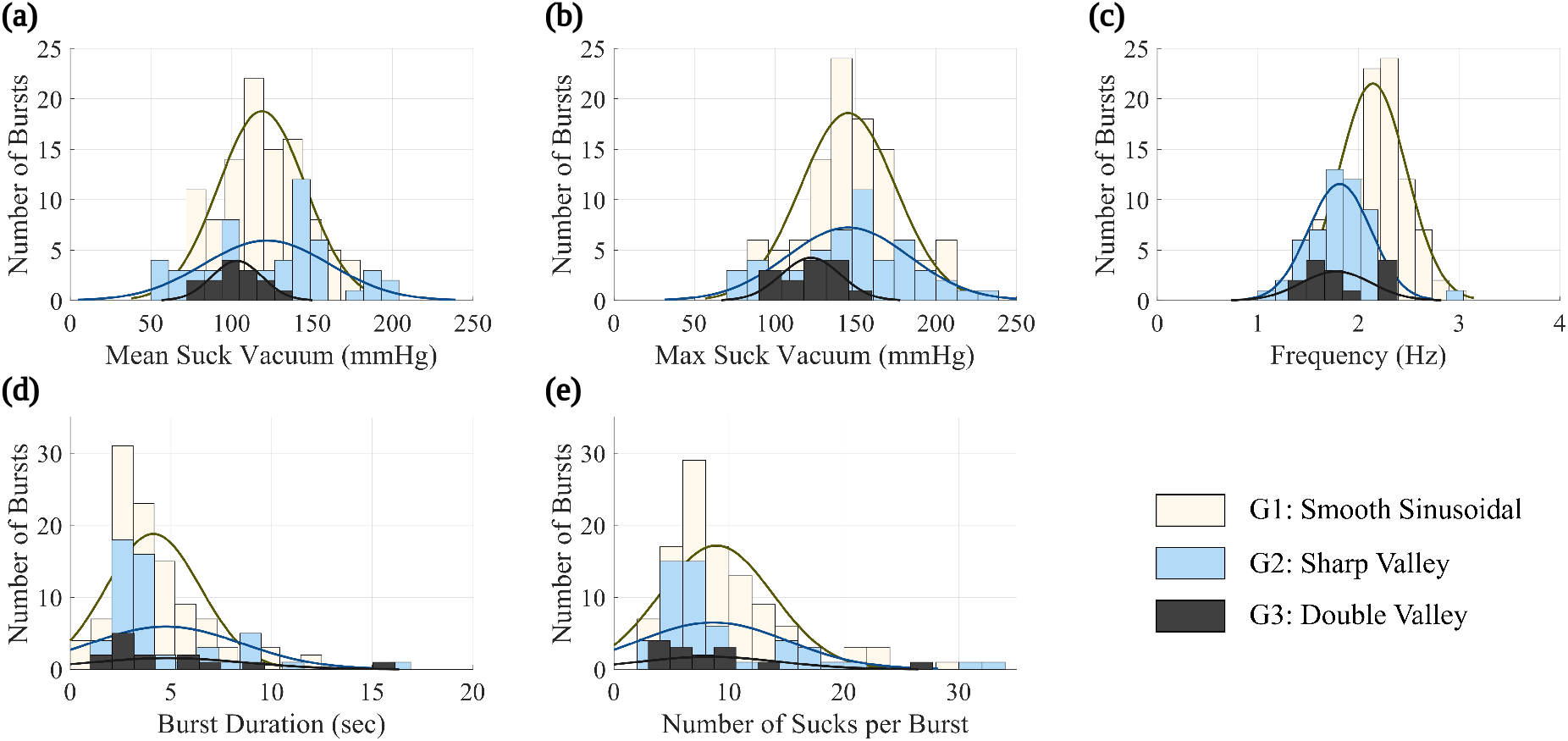
Distribution plots of the three classified groups based on signal shape: (a) Mean suck vacuum (b) Max suck vacuum (c) Frequency (d) Burst duration (e) Number of sucks per burst. The distributions of mean suck vacuum, max suck vacuum and frequency of the three groups were confirmed to be normally distributed using the Shapiro-Wilk normality test. The Welch’s t-test, which requires the data to be normally distributed, was then used to compare these parameters across the three groups for any statistical differences. Burst duration (d) and Number of sucks per burst (e) appear to be right-skewed distributions.

In our statistical analysis, classified the shape of the profiles into three main categories: smooth sinusoidal (18 neonates, 106 bursts), sharp valley (10 neonates, 53 bursts), and double-valley (2 neonates, 14 bursts). Histograms of the NNS parameters from the three groups are shown in Figure 6. It can be observed in Figure 6 (a-c) that the distributions of mean suck vacuum, max suck vacuum, and frequency are normally distributed. This was confirmed using the Shapiro-Wilk normality test. We then performed 2-sample Welch’s t-tests, which requires that data to be normally distributed, on the these parameters across the three groups to find any statistical differences between the groups. The results are shown in Table V. There were several statistically significant differences in mean suck vacuum, max suck vacuum and frequency between the three groups. There were no significant differences observed in burst duration and number of sucks per burst. These profile characteristics persist throughout the entire suckling signal of each infant. If the infant displays a signal shape corresponding to a sharp valley, we can observe this pattern throughout the entire suckling profile.

**TABLE V.** Welch’s t-test results comparing mean suck vacuum, max suck vacuum and frequency between the three groups of NNS profile shapes.

| Parameters | Groups | t | p |
| --- | --- | --- | --- |
| Mean Suck Vacuum (mmHg) | 1 vs. 2 | -0.49 | 0.62 |
|  | 1 vs. 3 | 3.16 | **0.005** |
|  | 2 vs. 3 | 2.72 | **0.010** |
| Max Suck Vacuum (mmHg) | 1 vs. 2 | -0.02 | 0.99 |
|  | 1 vs. 3 | 4.04 | **<0.001** |
|  | 2 vs. 3 | 3.22 | **0.003** |
| Frequency (Hz) | 1 vs. 2 | 6.15 | **<0.001** |
|  | 1 vs. 3 | 3.70 | **0.002** |
|  | 2 vs. 3 | 0.33 | 0.74 |

\* **Bold values are statistically significant.**

## IV. DISCUSSION

The results from our study show measurements in agreement with values reported in the literature. Our system demonstrates features and capabilities that addresses the clinical needs of an easy-to-use, accurate, and safe system. The immediate feedback of suckling performance allows clinicians to troubleshoot breastfeeding problems with greater accuracy using objective data.

Our region of interest analysis show differences in suckling profile shapes. These may relate to infant oral motor restrictions and function, and therefore are worthy of further study. A detailed burst analysis of the NNS data from the 30 neonates showed that there were statistically significant differences in key NNS parameters between neonates with different suckling profile shapes. These results suggest that the shape of the suckling profile can play an important role in evaluating the suckling mechanics of the infants. To our knowledge, this has not been reported in the literature. As more data from a larger population of neonates becomes available, we aim to further investigate the shape of the infant suckling profile as it relates to oral motor functions or disorders and also map key parameters of the profile, e.g., the sharpness of a given suckling vacuum event, to the severity of certain conditions.

The technical improvements that can be implemented in such a system include reducing the size of the data acquisition unit and incorporating Bluetooth capabilities to eliminate the cable to the computer. While this may further reduce the size of the system, it may also increase the system’s operating complexity and cost due to wireless pairing, data security considerations, and biosafety challenges caused by the proximity of the suckling unit to the electronics.

Clinical testing in this study examines a small sample size of infants and will expand further to investigate the system’s ability to capture profiles that reflect poor vacuum, coordination, fatigue, respiratory asynchrony, and varying maturation levels. More importantly, we aim to further investigate the shape of the infant suckling profile as it relates to oral motor functions or disorders. We hypothesize that existing systems have not yet demonstrated the subtle changes in the signal due to engineering design problems such as the use of large elastic tubing and the presences of large dead air volumes within the system that may dampen or reduce the sensitivity of the measurements.

Expression pressure is a common measurement capability of systems in the literature aimed at tracking premature infant oral motor feeding readiness. This typically occurs in bottle feeding. Our aim is to target breastfeeding, therefore, future iterations of our system may be modified for an infant’s suckling assessment at the breast. The pacifier can be removed and affixed to feeding tubes placed in the mouth while the baby is nursing. This permits comparison of non-nutritive and nutritive suckling skills. Such a system is reported in the literature by Chen et al. [16] and can be further investigated through larger studies with more infants in various clinical environments to better determine the feasibility of the feeding-tube system.

## V. CONCLUSIONS

In this paper, we report on the design of a non-nutritive suckling system. We demonstrated use and application of the system in a clinical environment: a specialist clinic and a general pediatric facility. Thirty neonates were enrolled in the study and their non-nutritive suckling profile was successfully recorded and analyzed in real time. The proposed system allows for objective measurements and quantitative analysis of an infant’s suckling profile. The system software interface automatically extracts features from the profile including the maximum and mean vacuum amplitude, suckling frequency, mean suck cycle, number of sucks, number of bursts, and the burst duration.

Like with all available systems and devices in the literature, the broader adoption of this technology in routine clinical practice will be a key challenge. Our future work will investigate the interpretation of these signals with respect to the norm (e.g., burst duration as it relates to endurance, maximum amplitude as it relates to suck vigor, etc.). As we collect more infant suckling profiles, this will enable us to establish a clear understanding of normal versus abnormal patterns of suckling, perhaps correlated to specific medical conditions at first identified by other means. These subtle suckling deviations can better distinguish infant-based interventions to optimize breast milk intake. Additionally, our study shows that real-time analysis feedback is important in the clinical environment, as measurements can be affected by infant behavior, preferences, and seal. With real-time data, repeating measurements as needed was crucial to obtaining and analyzing data in the clinic to ensure they had sufficient quality.

Ultimately, the challenge of diagnosing breastfeeding issues in mother-infant dyads remains a very complex and multidimensional problem. Our system aims to remove a facet of subjectivity in digital suckling examinations, by providing an objective quantification of suckling, working towards a clinical consensus within the medical and clinical community. This is with the overall goal of helping infants and mothers reach positive breastfeeding outcomes through referral and intervention pathways based on objective measurements. Extended applications of this system can include research of oral-motor or neurological development in infants, at-home intraoral vacuum monitoring system for infants, and as a rapid diagnostics tool in hospitals.

## Data Availability

All data produced in the present study are available upon reasonable request to the authors.

## ACKNOWLEDGMENTS

The authors are grateful to the University of California San Diego for provision of funds and facilities in support of this work via the Galvanizing Engineering in Medicine grant scheme and the Academic Senate. The authors are also grateful to the William H. and Mattie Wattis Harris Foundation for their kind financial support of this work. We are also grateful to the *Device Acceleration Center* of the Altman Clinical and Translational Research Institute for in-kind support of the clinical effort.

